# Enhancing evidence-based guidelines using trial emulation in electronic health records: Real-world effects of empagliflozin in people with type 2 diabetes

**DOI:** 10.1101/2025.09.17.25335885

**Authors:** David K Ryan, Ruth H Keogh, Elizabeth Williamson, R Thomas Lumbers, Karla Diaz-Ordaz, Anoop D Shah, Patrick Bidulka

**Affiliations:** Institute of Health Informatics, University College London, UK; Medical Statistics Department, London School of Hygiene and Tropical Medicine, UK; National Institute for Health and Care Research University College London Hospitals Biomedical Research Centre, University College London Hospitals NHS Foundation Trust, London, UK; Department of Statistical Science, University College London, UK; Department of Health Services Research and Policy, London School of Hygiene & Tropical Medicine, UK

**Keywords:** Diabetes Mellitus, Type 2, empagliflozin, Dipeptidyl-Peptidase IV Inhibitors, Electronic Health Records, Evidence-Based Medicine

## Abstract

**Background:** There is growing interest in widening the use of sodium-glucose co-transporter-2 inhibitors (SGLT2i) to all people with type 2 diabetes mellitus (T2DM). However, pivotal randomised controlled trials (RCTs) evaluated these drugs only in highly selected populations, often lacking generalisability to real-world populations. Understanding the effects of SGLT2i in populations, where RCT evidence may be lacking is essential to help inform guideline development. To address this, we estimated the effect of empagliflozin in a real-world users, many of whom would not have been eligible for the pivotal EMPA-REG RCT.

**Methods:** We designed a trial emulation in UK primary care data, based on the EMPA-REG RCT, to assess the effect of empagliflozin in a more clinically relevant population. Adults with T2DM initiating empagliflozin (intervention) or dipeptidyl peptidase-4 inhibitors (active control) between 1 Jan 2014–31 Dec 2022 were included. Eligibility was extended to both RCT-eligible and RCT-ineligible individuals. The effect of empagliflozin on all-cause mortality was estimated using an adjusted Cox proportional hazards model, with stratified analyses by RCT eligibility.

**Findings:** The majority of people prescribed empagliflozin would not have met the EMPA-REG RCT eligibility criteria (11011/13239, 83.2% RCT-ineligible). During follow-up, all-cause mortality occurred in 551/13239 (4.2%) of the empagliflozin group and 6589/49264 (13.4%) of the active control group (adjusted HR 0.76, 95% CI 0.69–0.83). There was no evidence of differential treatment effect by RCT eligibility status (p-interaction=0.27).

**Interpretation:** Patients prescribed empagliflozin in real-world settings differ substantially from those enrolled in the EMPA-REG RCT. Using electronic health records, we demonstrate that the mortality benefit observed in EMPA-REG extends to a broader, more diverse real-world population, including those excluded from the original RCT. These findings provide a novel source of real-world evidence supporting the wider use of empagliflozin in routine clinical practice.

**Funding:** DKR, NIHR Doctoral Fellow (NIHR304689), is funded by the NIHR for this research project. RHK is funded by UK Research and Innovation (Future Leaders Fellowship). EW is funded by a Wellcome Senior Research Fellowship (grant number 224485/Z/21/Z). RTL is supported by the National Institute for Health Research University College London Hospitals Biomedical Research Centre, the UCL British Heart Foundation Accelerator (AA/18/6/34223), the MRC/NIHR Rare Disease Research UK Cardiovascular Initiative.

**Evidence before this study:**

- Randomised controlled trials (RCTs), including the seminal EMPA-REG RCT, have shown that empagliflozin, a sodium-glucose co-transporter-2 inhibitor (SGLT2i), significantly reduces mortality in people with type 2 diabetes (T2DM) and established atherosclerotic cardiovascular disease.
- There is growing interest in expanding SGLT2i use to broader populations, with draft UK National Institute for Health and Care Excellence (NICE) 2025 guidelines proposing it as first-line therapy for all people with T2DM, in combination with metformin.
- Given the growing divergence between RCT evidence and proposed universal use of SGLT2i in T2DM management, the real-world effectiveness of empagliflozin, particularly among patients under-represented in RCTs, remains uncertain.

**Added value of this study:**

- Using the trial emulation framework in UK primary care data, we demonstrate that the population receiving empagliflozin in current practice already differs substantially to the population enrolled in the EMPA-REG RCT.
- We demonstrate that relaxing the stringent RCT eligibility criteria imposed by EMPA-REG RCT did not alter the treatment effect in real-world populations, with strong evidence of a mortality benefit in people, regardless of RCT eligibility status.

**Implications of all the available evidence:**

- These results support broader use of empagliflozin in T2DM management, beyond the eligibility criteria of landmark RCTs.
- These findings provide critical real-world evidence supporting a universal SGLT2i strategy in T2DM management, which is of direct relevance to current guideline deliberations.

## INTRODUCTION

Type 2 diabetes mellitus (T2DM) is a growing global health concern. In the UK, over 3.5 million people live with T2DM and by 2040, the prevalence of T2DM is estimated to increase by 50%.^1^ Since 2022, UK National Institute for Health and Care Excellence (NICE) treatment guidelines for T2DM recommend metformin and sodium-glucose cotransporter-2 inhibitors (SGLT2i) as dual first-line agents for people with T2DM and concomitant cardiovascular disease (CVD), risk factors for CVD or heart failure.^2^ The 2022 NICE guideline committee decision was based on extrapolations from randomised controlled trials (RCTs), such as the EMPA-REG RCT, which showed significant cardioprotective and mortality benefits of SGLT2i.^2^ This follows similar recommendations by the European Society of Cardiology,^3^ European Association for the study of Diabetes and the American Diabetes Association.^4^

In late August 2025, the UK National Institute for Health and Care Excellence (NICE) released a draft updated guideline for the management of type 2 diabetes mellitus (T2DM) for public consultation.^5^ A key proposal in this draft is to recommend sodium-glucose co-transporter-2 inhibitors (SGLT2i) alongside metformin as first-line agent for all people with T2DM. This is based on evidence from randomised controlled trials (RCTs) and network meta-analyses showing that combined therapy was more effective than metformin monotherapy alone.^5^ If adopted, these guidelines would mark the most significant shift in T2DM care in a decade, with major implications for patients, clinicians and wider society. They also would mark a further extrapolation of clinical practice from RCTs, where the evidence base for SGLT2i was established in highly selected populations of people with T2DM.

EMPA-REG RCT was a large multi-centre trial which randomised 7020 people with T2DM and established CVD to receive either the SGLT2i, empagliflozin, or placebo.^6^ To be eligible for this study, people had to meet stringent eligibility requirements. For example, participants were required established CVD such as a previous myocardial infarction or unstable angina with evidence of coronary artery disease on coronary angiogram. This focus on participants with high CVD burden was consistent across other major SGLT2i trials.^7,8^ However, NICE and other bodies subsequently opted to recommend SGLT2i for people with T2DM who did not have established CVD, such as a previous cardiac event, but were considered at high-risk for CVD.^2^ Given the growing divergence between RCT evidence and proposed universal use of SGLT2i, the real-world effectiveness of empagliflozin, particularly among people under-represented in RCTs, remains uncertain.

There is increasing interest in using observational data to generate real-world evidence for treatments, either to complement existing research or fill gaps in the current knowledge base.^9,10^ In addition, real-world evidence can give insights that may not be studied, or indeed feasible to study, through RCTs alone – such as long-term effects of medications, treatment effects in trial-excluded populations, heterogeneous treatment effects as well as comparative effectiveness and safety studies, among others. ^9,10^ In the context of guideline development, real-world evidence can play a particularly to help inform decisions, where evidence from RCT and associated meta-analyses may be lacking or limited.

Using a trial emulation framework, we leverage UK primary care data to estimate the real-world treatment effect of a key anti-diabetes drug in a broader population compared to those enrolled in RCTs. In this way, we derive evidence for SGLT2i in wider populations, which may be useful to inform current guideline deliberations that affect all people living with T2DM. In our emulation, we modified the design of the EMPA-REG RCT to study the real-world treatment effect of empagliflozin in broader populations. We subsequently estimate the treatment effect of empagliflozin among initiators who would have been excluded for the EMPA-REG RCT, thereby generating evidence for empagliflozin for the large population currently under-represented in pivotal RCTs.

## METHODS

### Data sources

We used de-identified UK primary care electronic health record data from The Health Improvement Network (THIN), a Cegedim database. THIN contains data on demographics, lifestyle, diagnoses, prescriptions, examinations, laboratory results, and clinical measurements (blood pressure, BMI).

Diagnoses are recorded using Read codes, a standardised terminology. The database is representative of the UK population.^11^

### Study design

We designed a trial emulation adapting the EMPA-REG RCT. Eligibility criteria were relaxed to capture the broader population of real-world empagliflozin initiators and better reflect clinical practice. The study followed best-practice guidelines for trial emulation,^12^ as described in table 1. A new-user active comparator design compared mortality between initiators of empagliflozin and dipeptidyl peptidase-4 inhibitors (DPP-4i: alogliptin, linagliptin, sitagliptin, saxagliptin, vildagliptin). DPP-4i drugs were selected as the active comparator as they have no known cardioprotective or mortality benefit.^13^

**Table 1:** Study design of the RCT and trial emulation Table highlighting the main design elements of the trial emulation, with corresponding design of the EMPA-REG randomised controlled trial (RCT). T2DM: type 2 diabetes mellitus; SGLT2: sodium glucose co-transporter 2 inhibitor (empagliflozin, dapagliflozin, canagliflozin and ertugliflozin); DPP-4i: dipeptidyl peptidase-4 inhibitors (alogliptin, linagliptin, sitagliptin, saxagliptin and vildagliptin); BMI: body mass index; HbA1c: glycated haemoglobin A1c.

|  | **EMPA-REG RCT** | **Trial emulation** |
| --- | --- | --- |
| **Inclusion criteria** | Individuals with T2DM, aged over 18 years, with established coronary artery disease (previous acute coronary syndrome, established coronary artery disease), stable glycaemic control and BMI $\leq 45$ kg/m<sup>2</sup> | Individuals with T2DM, aged over 18 years, who are SGLT2i and DPP-4i naïve and have an incident prescription for either empagliflozin or DPP-4i between 1<sup>st</sup> January 2014 – 31<sup>st</sup> December 2022 |
| **Exclusion criteria** | People with baseline renal impairment (eGFR < 30 mL/min/1.73m<sup>2</sup>), recent acute coronary syndrome, stroke or transient ischaemic attack, liver disease, recent pregnancy among other factors. See supplementary material for full list | Past medical history of pancreatitis or ketoacidosis |
| **Treatment strategies** | Empagliflozin or placebo | Incident initiators of either empagliflozin or DPP-4i (active comparator design) |
| **Treatment assignment** | Randomisation with analysis via intention-to-treat | Analysed as observational analogue of intention-to-treat |
| **Day zero** | Date of randomisation following a two-week open-label placebo run-in period | Date of first incident prescription for either empagliflozin or DPP-4i between 1<sup>st</sup> January 2014 – 31<sup>st</sup> December 2022, whichever occurred first |
| **Follow-up period** | Median follow-up of 3.1 years | Follow-up begins at the day zero and ends at first of: date of death, date of leaving general practice or 31<sup>st</sup> December 2023 |
| **Confounders** | Adjusted for chance baseline differences between groups (age, gender, baseline BMI, baseline HbA1c, baseline eGFR, geographical region) | Adjusted for confounding variables at baseline in an adjusted Cox proportional hazards model |
| **Outcomes** | Primary outcome: time-to-first composite outcome event (cardiovascular death, non-fatal stroke, non-fatal myocardial infarction)<br>Secondary outcome: time-to all-cause mortality | Primary outcome: time-to all-cause mortality |
| **Causal treatment effect** | Adjusted hazard ratios (HR) for composite outcome and all-cause mortality, both estimated by a Cox proportional hazard models | Adjusted HR for all-cause mortality estimated via a Cox proportional hazards model |

Trial emulation is a methodological approach that replicates either an existing or a hypothetical clinical trial in an observational setting.^12,14^ The purpose of the trial emulation framework is to reduce bias in observational studies by pre-specifying eligibility criteria, time zero, treatment strategies, follow-up and outcome measures, and is advocated by the UK National Institute for Health and care Excellence.^10^ A key benefit of targeting an existing RCT is that the observational results can be ‘benchmarked’ to the published results of the RCT, giving confidence that the real-world analysis are accounting for confounding appropriately. This can then support the emulation of hypothetical or modified trials, allowing us to draw meaningful inferences about causal effects of medications in real-world practice.

Our trial emulation was primarily a hypothetical trial of empagliflozin in routine practice, informed by EMPA-REG but in a broader population. We also compared treatment effects in RCT-eligible and RCT-ineligible initiators of empagliflozin, enabling benchmarking against EMPA-REG results.

### Study population

The trial emulation analysed data from adults with T2DM who had an incident prescription for either empagliflozin or an active control, DPP-4i between 1^st^ January 2014 – 31^st^ December 2022. These study dates were chosen to align with the introduction of SGLT2i in the UK and the subsequent updates to NICE guidelines in June 2022, which thereafter prioritised SGLT2i as an oral anti-diabetic agent.^2^ People who had prior use of SGLT2i or DPP-4i before cohort entry date were excluded to avoid dilution of treatment effect from a drug-class effect. People were analysed according to their treatment assignment on day zero in an intention-to-treat manner, regardless of post-baseline changes in treatment assignment (e.g. discontinuation, switching or intensification).

Unlike the stringent eligibility criteria defined in the EMPA-REG RCT,[5] we did not exclude individuals based on criteria such as: presence of established atherosclerotic disease, glycated haemoglobin A1c (HbA1c), body mass index (BMI) or renal function thresholds. The only inclusion criteria we applied was being aged over 18 and a confirmed diagnosis of T2DM. People were excluded from the trial emulation if they had a GP registration date within a year of cohort entry to ensure sufficient capture of baseline covariates. People were also excluded if they had a history of pancreatitis or ketoacidosis because they would have a low chance of being prescribed an SGLT2i due to clinical recommendations advising caution in prescribing SGLT2i to such individuals.^15^

### Outcomes

The primary outcome in the EMPA-REG RCT was a composite three-point major adverse cardiovascular event, consisting of CVD death, non-fatal myocardial infarction and non-fatal stroke.^6^ However, cause-specific mortality data was not available in the THIN database. As a result, the primary outcome for the trial emulation was all-cause mortality. This was reported as a secondary outcome in the EMPA-REG RCT, allowing for comparison of results. Date of death was estimated using a validated algorithm,^16^ which gathers various recorded death codes from primary care records and integrates the data to determine the most accurate estimated date of death. Follow-up begins at the date of first prescription of empagliflozin or DPP-4i, and ends at first of: date of death, date of leaving general practice or 31^st^ December 2023.

### Confounders

Confounders were identified based on clinical knowledge, and included age, sex, ethnicity, socioeconomic status (defined by the index of multiple deprivation quintile at the level of a GP practice), calendar year of cohort entry, comorbidities, co-prescribed medication and laboratory and clinical measurement values (blood pressure, BMI, HbA1c, cholesterol) among others. These are summarised in a directed acyclic graph (supplementary material 1). Pre-existing published codelists from the Health Data Research UK phenotype library were used to define covariates. Details and codelists for covariates are described in the supplement (supplementary material 2 – 3). Ethnicity was re-categorised according to the most recent UK census classification (supplementary material 3).[22]

Glycated haemoglobin A1c (HbA1c) was defined based on the most recent value up to 180 days before time zero. This 180-day window was selected as NICE recommend that HbA1c is measured every 6 months in people with T2DM.^17^ For other measurements such as systolic blood pressure, BMI, cholesterol and eGFR, the most recent value within a window of 540 days before baseline was used. This was informed by the UK primary care Quality and Outcomes Framework, which recommend patients with T2DM have a full clinical review annually, with additional time allocated for delays and data entry.^18^ If values were not recorded, or not available within the eligible window for the covariate in question, the value was defined as missing. Further details are available in supplementary material 2. Other covariates were defined based on the most recently available information at day zero.

### Randomised controlled trial eligibility status for empagliflozin users

Once our trial emulation population was defined, we assessed what proportion of these people would have met the stringent eligibility criteria of the EMPA-REG RCT. The mapping of the RCT eligibility criteria to the observational setting is described in supplementary material 5. This enabled us to define RCT eligibility status for each user of empagliflozin or DPP-4i, based on whether they would have met the EMPA-REG RCT criteria. Several RCT eligibility criteria related to variables that had incomplete data capture such as laboratory tests at baseline (e.g. estimated glomerular filtration rate, baseline HbA1c). People were defined as RCT eligible if these values were missing or not measured within the pre-defined window before cohort entry date.

### Statistical analysis

Full details are in the supplementary appendix. Demographics, clinical measurements, prescribed medication and comorbidities were described for the trial emulation population overall, and by treatment group, and were compared to the published table of subject characteristics from the EMPA-REG RCT.

An adjusted Cox proportional hazard model was used to estimate the hazard ratio for all-cause mortality, with time on study as the timescale. The Cox proportional hazards model was adjusted for confounders that are defined in the study directed acyclic graph. BMI, LDL and HDL cholesterol, eGFR, HbA1c, systolic blood pressure, ethnicity, and smoking status had missing data. Missing values were handled using multiple imputation by chained equations (MICE),^19^ generating five fully imputed datasets with pooled estimates derived using Rubin’s rules.

Three-year risk difference in all-cause mortality between treatment groups and associated number needed to treat (NNT) to prevent one death were estimated using a g-computation approach, with confidence intervals estimated using a bootstrap imputation procedure.^20^ Three years of follow-up was selected as this matches the median follow-up period of the EMPA-REG RCT.[5] For sensitivity analysis, an inverse-probability of treatment weighted (IPTW) Cox proportional hazard model was employed to estimate the average treatment effect, and also the average treatment effect on the treated.

We then estimated the real-world treatment effect of empagliflozin, stratified according to RCT eligibility status, by adding an interaction term to the adjusted Cox proportional hazards model. This tested whether treatment effect differed by RCT eligibility status and provided a hazard ratio (HR) for the effect of empagliflozin vs. DPP-4i in both RCT ineligible and RCT eligible initiators of empagliflozin.

Results from the RCT eligible subgroup enabled benchmarking against EMPA-REG RCT estimates using pre-defined agreement metrics. These metrics are consistent with other trial emulations:^21^

1. Statistical significance agreement: Emulated estimates and confidence intervals align on the same side of the null as the RCT
2. Estimate agreement: Emulated estimates fall within the 95% confidence interval of the RCT estimate
3. Standardised difference: Compares the difference in effect size between the RCT and trial emulation, allowing formal hypothesis

### Ethical approval

This study was approved by the THIN scientific review committee (SRC 23-010, Dec 2023) and the London School of Hygiene and Tropical Medicine research ethics committee (reference: 29647). THIN has database-wide ethics approval from the South Central - Oxford C Research Ethics Committee (20/SC/0011).

## RESULTS

### Trial emulation population and baseline characteristics

A total of 62503 people with T2DM initiated either empagliflozin (N=13239, 21.2%) or DPP-4i (N=49264, 78.8%) between 1^st^ January 2014 – 31^st^ December 2022. A total of 7140 deaths (11.4%) were recorded in this trial emulation. Death occurred in 551/13239 (4.2%) of the empagliflozin group and in 6589/49264 (13.4%) of the DPP-4i (active control) group. The number of person-years-at-risk were 200,646 years and the longest duration of follow-up was 9.6 years.

In the trial emulation population, 12970 individuals (20.8%) would have met the eligibility criteria for the EMPA-REG RCT (5). A total of 13239 patients with T2DM received a first prescription of empagliflozin within the study period, of whom 83.2% (N=11011/13239) would not have met the eligibility criteria for the EMPA-REG RCT. Most initiators of study drugs would have been excluded for not having established CVD at baseline (excluded N = 43672, 70.0%) or having a recorded baseline HbA1c outside of the appropriate range (excluded N = 14953, 23.9%). There were 36088 individuals (57.7% of trial emulation population) defined as RCT ineligible based on having a single exclusion criterion, with a further 13445 people (21.5%) defined as RCT ineligible based on having more than one exclusion factor.

The trial emulation population was older (average age in trial emulation 64.0 years, standard deviation [SD] 13.5 vs. average age in the RCT 63.1, SD 8.6, table 2), had substantially greater representation of women (42.5% of the population were female in the trial emulation vs. 28% in the RCT, table 2) and had a higher baseline HbA1c compared to the RCT population (average baseline HbA1c in trial emulation: 74.3mmol/mol, SD 17.8 vs. 64.7mmol/mol in the RCT, table 2).

**Table 2:**
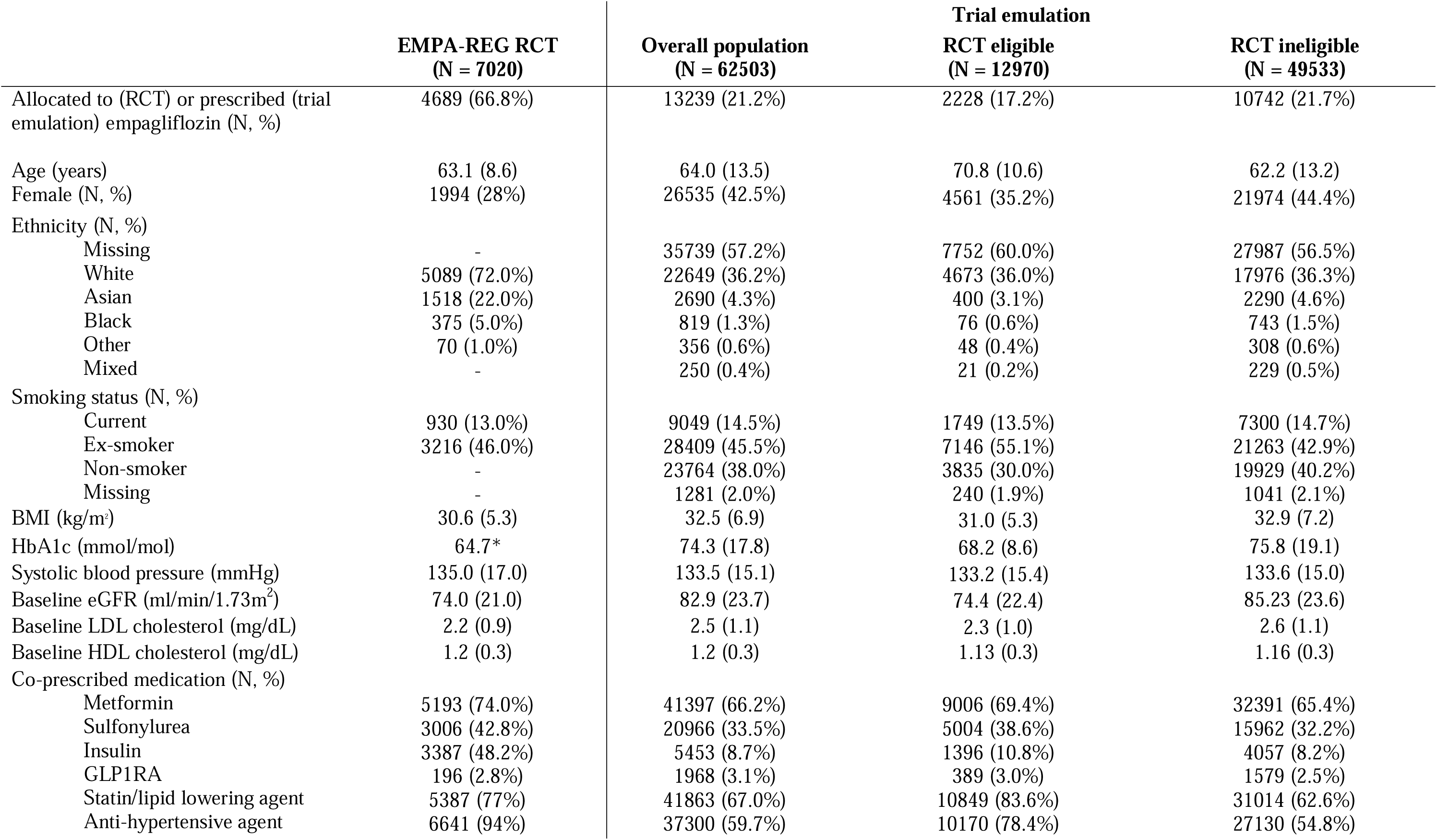
Demographics of the RCT and trial emulation populations, with further stratification according to RCT eligibility status in real-world population Table comparing the randomised controlled trial (RCT) and the trial emulation population, with further stratification by RCT eligibility status. Data presented as mean (standard deviation) or count (column percentage) for consistency with trial reporting. *Standard deviation not provided for HbA1c mmol/mol unit. BMI: body mass index, HbA1c: glycated haemoglobin A1c, eGFR: estimated glomerular filtration rate, LDL: low density lipoprotein, HDL: high density lipoprotein, GLP1RA: GLP-1 receptor agonist.

The trial emulation cohort had a lower burden of co-prescribed medication compared to the RCT population (table 2). For example, much greater numbers were co-prescribed insulin in the RCT compared to the trial emulation (3387 people in the RCT, 48.2% of RCT cohort vs. 5453 people in the trial emulation, 8.7%). Similar patterns can be seen across most anti-diabetes drug classes and anti-hypertensive agents. This likely reflects different burdens of diseases between RCT and real-world populations, but also geographical variations in clinical practice.

As expected in a RCT, the empagliflozin and control groups are approximately balanced for all key characteristics. However, in the trial emulation population, people who commenced empagliflozin tended to be younger but have worse metabolic markers (higher average BMI and higher average HbA1c) compared to people initiated on DPP-4i (supplementary material 8). People in the empagliflozin group tended to have lower burden of comorbidities – particularly for cardiovascular disease (present in 3424/13239, 25.7% of the empagliflozin group vs. 15407/49264, 31.3% of the DPP-4i group), and dementia (present in 50/13239, 0.4% of the empagliflozin group vs. 1172, 2.4% of the DPP-4i group). In addition, people in the empagliflozin were less likely to be prescribed metformin, sulfonylurea, anti-hypertensives and lipid lowering drugs (supplementary material 8).

### Trial emulation outcome analysis

In the trial emulation, the primary outcome of all-cause mortality occurred in 551/13239 (4.2%) of the empagliflozin group and in 6589/49264 (13.4%) of the active control group. The adjusted hazard ratio for all-cause mortality in the Cox proportional hazards model was 0.76 (95% CI 0.69 – 0.83), providing strong evidence of a mortality benefit for people initiated on empagliflozin compared to those who initiated DPP-4i (figure 2).

**Figure 1:**
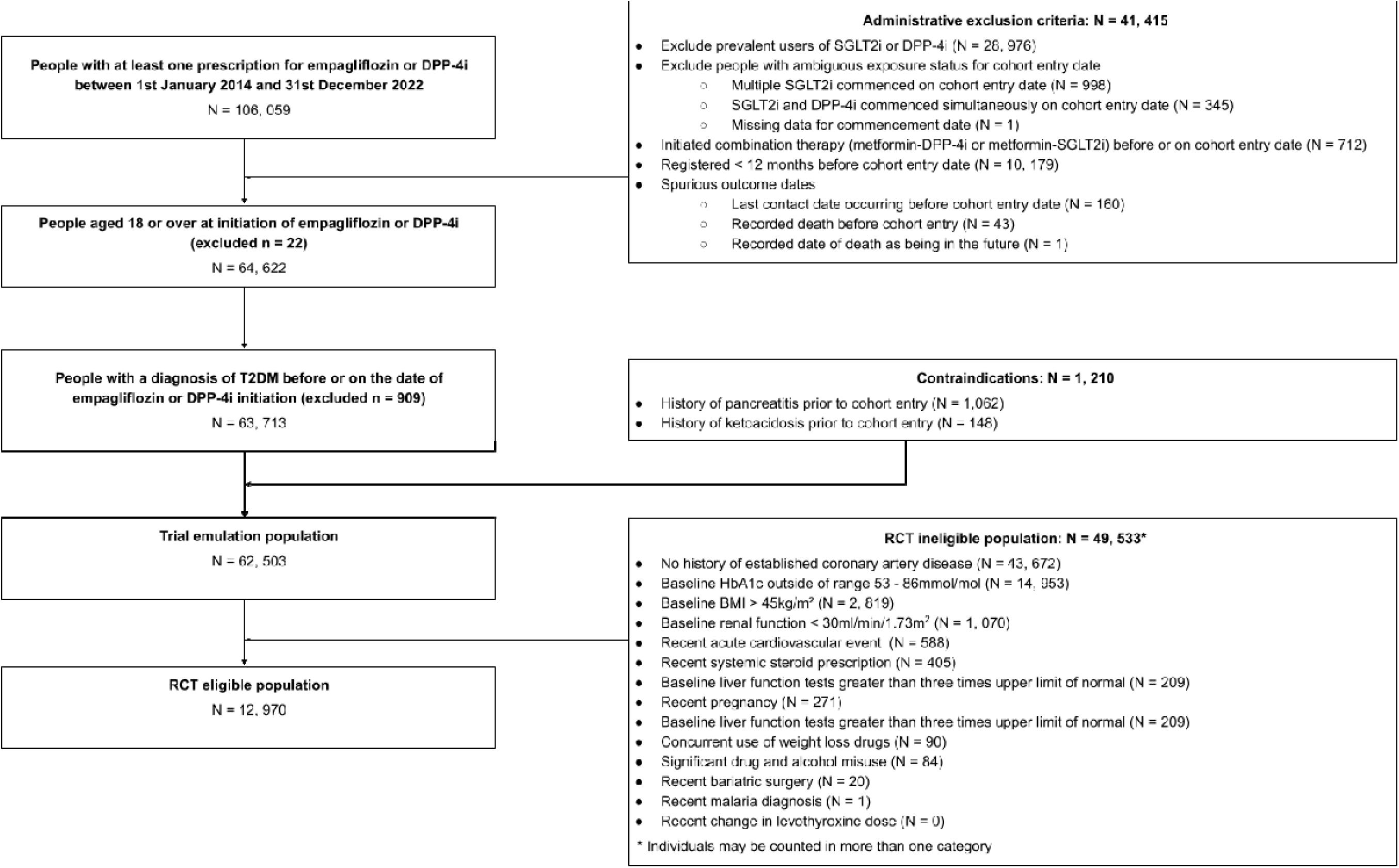
Flow diagram for the trial emulation Flow diagram showing attrition of people according to eligibility criteria for the trial emulation. The trial emulation is a modified design of the EMPA-REG RCT and relaxes the stringent eligibility criteria of the RCT. SGLT2: sodium glucose co-transporter 2 inhibitor: empagliflozin, dapagliflozin, canagliflozin, ertugliflozin. DPP-4i: dipeptid peptidase-4 inhibitors (alogliptin, linagliptin, sitagliptin, saxagliptin, vildagliptin); T2DM: type 2 diabetes mellitus; HbA1c: glycated haemoglobin A1c; BMI: body mass index.

**Figure 2:**
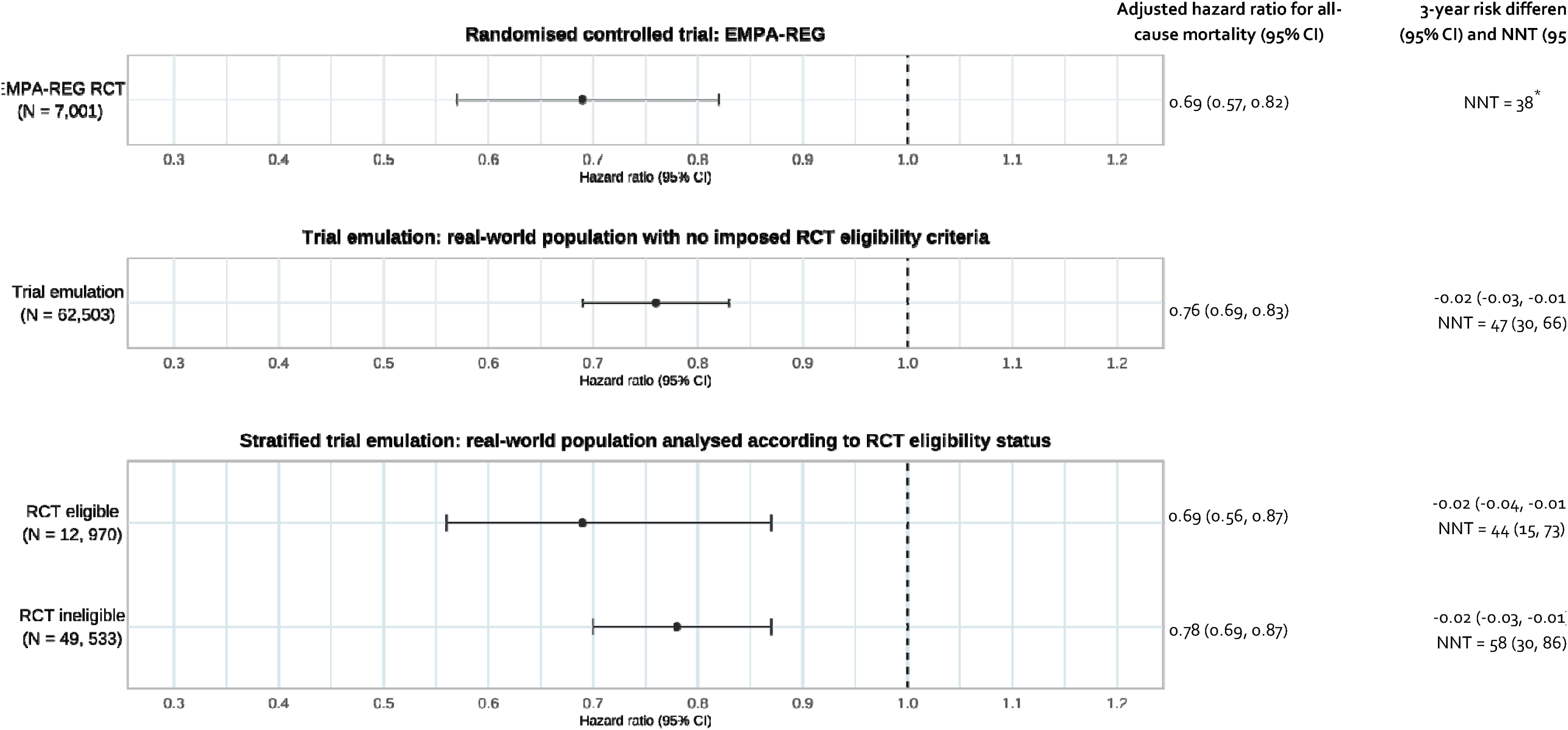
Results showing the adjusted hazard ratio for all-cause mortality in the RCT and trial emulation Forest plot showing the hazard ratio and 95% confidence intervals (CI) for all-cause mortality, 3-year risk difference and number needed to treat (NNT) to prevent on death in the randomised controlled trial (RCT), trial emulation and stratified trial emulation analysis. The trial emulation population represents initiators of study drugs (empagliflozin or DPP-4i), without imposing RCT-defined eligibility criteria. The stratified trial emulation refers to an adjusted Cox proportional hazards model where there is a additional interaction between RCT eligibility status and empagliflozin, allowing estimation of the hazard ratio for empagliflozin vs. DPP-4i in both RCT eligible and RCT inelig real-world populations. * Risk difference and 95% CI not published in post-hoc analysis of the EMPA-REG RCT.

There was reduced three-year risk of mortality in the empagliflozin group compared to the DPP-4i group (risk difference −0.02, 95% CI −0.03, −0.01, figure 2). The corresponding NNT is 47 people (95% CI 30, 66, figure 2). The IPTW analysis yielded similar results (supplementary material 11) for both the estimate as an average treatment effect and average treatment effect in the treated. These results were robust to the presence of unmeasured confounding in E-value analysis (supplementary material).

### Stratified trial emulation to assess treatment effect by RCT eligibility

Hazard ratios for all-cause mortality were consistent between RCT eligible and RCT ineligible initiators of empagliflozin (figure 2), with no evidence of an interaction between the treatment effect of empagliflozin and RCT eligibility status (p-value for interaction 0.27). The RCT-eligible strata-specific hazard ratios met all predefined agreement criteria with the published RCT results, confirming the successful benchmarking of the observational findings against the RCT findings.

The NNT at 3 years from the RCT was within the 95% confidence intervals for all trial emulation estimates, including both RCT eligible and RCT ineligible populations (figure 2).

## DISCUSSION

In this study, we applied a trial emulation framework to investigate the real-world treatment effect of empagliflozin in people with T2DM. Our findings confirm that the mortality benefits of empagliflozin observed in the EMPA-REG RCT are realised in real-world practice. Notably, we found that the majority of real-world empagliflozin users would not have met the eligibility criteria imposed by the EMPA-REG RCT. By modifying the design of the EMPA-REG RCT to reflect contemporary real-world utilisation, we demonstrate that individuals excluded from the RCT, but now receiving treatment with empagliflozin, have comparable treatment effect to those who were represented in the RCT. This supports the wider use of these drugs beyond the limited scope of utilisation within RCTs. It directly addresses a gap in evidence that is useful to current NICE deliberations around implementing a universal SGLT2i policy in T2DM management, particularly in populations where RCT evidence is lacking.

It is well-recognised that RCTs often have limited external generalisability, often due to strict eligibility criteria.^22^ A key finding of the present study is that as few as 16.8% of individuals with T2DM in the UK initiated on empagliflozin would have met the original EMPA-REG RCT eligibility criteria. This aligns with findings from a Taiwanese study, where only 18.7% of SGLT2i users met relevant RCT eligibility criteria.^23^

Our real-world evidence demonstrates consistent mortality benefits across both RCT eligible and ineligible real-world populations, many of whom have low burden, or absence of, CVD. This is a clinically important finding as it addresses a gap in the current evidence-base for empagliflozin, identified by both NICE committee deliberations and others.^2,5^ A previous meta-analysis of 7 RCTs (N = 4, 495) studying the effects of empagliflozin vs. placebo in people with T2DM with low-medium burden of CVD concluded that there was no evidence supporting significant reduction in all-cause mortality (HR 0.67, 95% CI 0.28 – 1.63).^24^ We, however, show a significant, and long-term mortality benefit in a diverse and clinically relevant population of people with T2DM. The difference between these findings and the present study likely reflects the larger population studied in the real-world analysis, providing greater precision.

## Strengths and limitations of this study

This study uses a large, representative UK primary care database, benefiting from comprehensive health record capture. Given that most diabetes care occurs in primary care settings with incentives for recording key covariates, data completeness is high. The study period (2014–2022) coincided with variable prescribing practices due to a lack of clear oral anti-diabetic guidelines, helping to reduce systematic bias between groups.^18^

A key aspect of the study design was the emulation of the EMPA-REG RCT in UK primary care data. To our knowledge, this is the first such emulation in a European setting. We were able to successfully benchmark the results of our real-world analysis to the published results of the RCT, giving us confidence that our analysis is appropriately handling confounding. In addition, this design helps avoid issues in other similar observational studies of T2DM medication such as immortal time bias and time-lag bias.^9,14^ We acknowledge the potential for residual confounding, for example by unmeasured covariates such as frailty. However, we do not believe that the presence of unmeasured confounding would be so substantial as to alter the conclusions of this study because of our rigorous modelling strategy, close replication of the existing RCT findings, and supportive quantitative bias assessment (E-value analysis, supplementary material). We also acknowledge that the primary outcome of the trial emulation is a secondary outcome of the EMPA-REG RCT, but cause-specific mortality data was not available within the electronic health record database.

This trial emulation did not assess safety outcomes, an important area for future research. Real-world safety signals may differ from RCT findings due to greater comorbidities and polypharmacy, increasing the risk of drug interactions and adverse effects.[34]

## CONCLUSIONS

The mortality benefit of empagliflozin, originally seen in the EMPA-REG RCT, is also observed in real-world settings, among a broader, more clinically relevant population of people with T2DM. This study provides robust real-world evidence to support the wider utilisation of empagliflozin in T2DM management, beyond the narrow eligibility criteria imposed by RCTs. This is of importance given the proposal to utilise SGLT2i as a first-line therapy for all people living with T2DM, extending the use of this therapeutic far beyond the initial scope of existing RCTs.

## Data Availability

Data is not publicly available but applications from eligible researchers can be made to the THIN scientific committee.

## ACKNOWLEDGEMENTS

The authors would like to acknowledge funding provided by the National Institute for Health and Care Research (NIHR) as well as the assistance of the local database manager at the University College London Institute of Health Informatics, Muhammad Qummer Ul Arfeen.

## AUTHORS’ CONTRIBUTIONS

- **David K Ryan**: Conceptualisation, methodology, formal analysis, visualisation, writing – original draft, writing – review & editing.
- **Ruth H Keogh**: Conceptualisation, methodology, formal analysis, supervision, writing – review & editing.
- **Elizabeth Williamson**: Methodology, supervision, validation, writing – review & editing.
- **R Thomas Lumbers**: Conceptualisation, data interpretation, clinical expertise, writing – review & editing.
- **Karla Diaz-Ordaz**: Methodology, writing – review & editing.
- **Anoop D Shah**: Conceptualisation, data curation, data analysis, supervision, funding acquisition, project administration, writing – review & editing.
- **Patrick Bidulka**: Conceptualisation, data curation, software, methodology, analysis, supervision, writing – review & editing.

## DATA SHARING AGREEMENT

Data cannot be shared publicly because of patient confidentiality; the study uses individual patient electronic health record data. Data are available from The Health Improvement Network (THIN) for researchers who meet the criteria for access to confidential data (contact via).

