## supplement for "Enhancing evidence-based guidelines using trial emulation in electronic health records: Real-world effects of empagliflozin in people with type 2 diabetes"

**Table of Contents**

***1.***	***Study directed acyclic graph***

***2.***	***Covariate definitions***

***3.***	***Ethnicity***

***4.***	***Dataset construction***

***5.***	***Mapping the EMPA-REG RCT eligibility criteria to real-world populations***

***6.***	***Statistical methods***

***7.	Missing data***

***8.***	***Demographics according to treatment randomisation/allocation***

***9.***  	***Full results table***

***10.***  	***E-value: quantitative bias assessment results***

***11.***  	***Covariate balance for the IPTW analysis***

***12.***  	***ATE and ATT results***

1.     	Study directed acyclic graph

Confounding factors were selected based on clinical domain knowledge and covariates adjusted in similar studies. This is summarised in a directed acyclic graph (DAG). Empagliflozin/DPP-4i is the exposure, with all-cause mortality being the outcome. Red circles denote confounders. eGFR: estimated glomerular filtration rate, HbA1c: glycated haemoglobin A1c, BMI: body mass index, SBP: systolic blood pressure. DAG was constructed using the online daggity tool ([[2]](https://www.dagitty.net/)).

**Supplementary figure 1:** Study directed acyclic graph

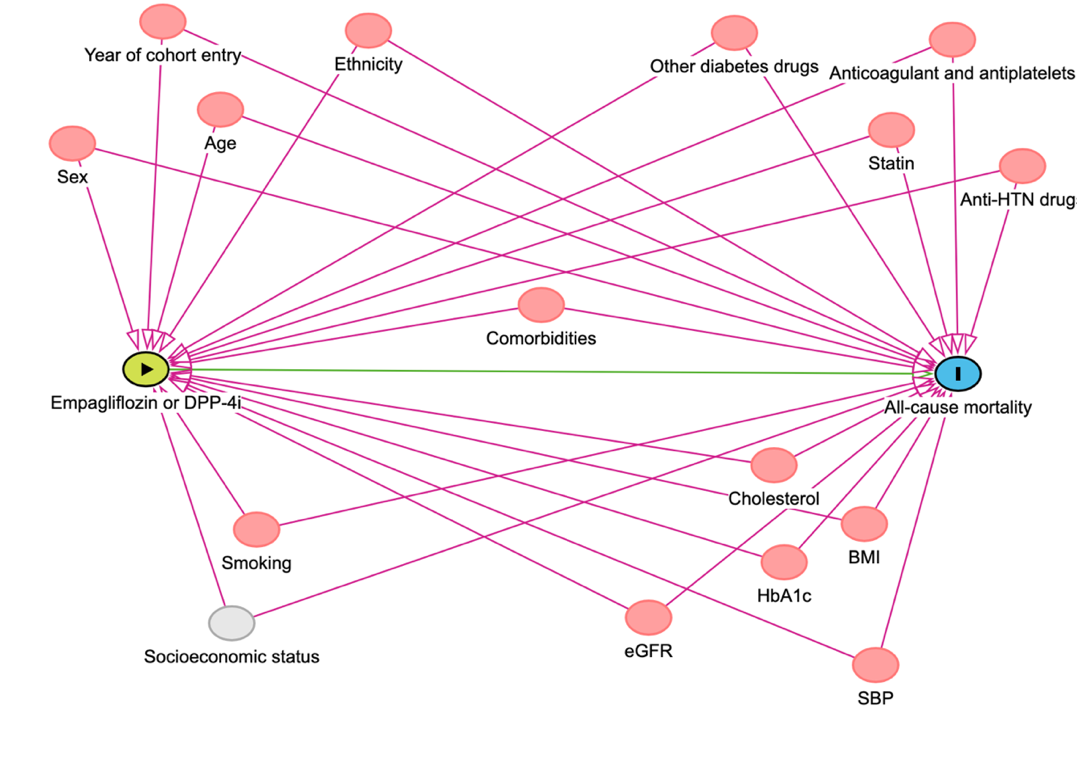

***Supplementary figure 1:*** *Figure showing confounding structures. DPP-4i: dipeptidyl peptidase-4 inhibitor, SBP: systolic blood pressure, eGFR: estimated glomerular filtration rate, HbA1c: glycated haemoglobin A1c, BMI: body mass index, Anti-HTN: anti-hypertensive agent (angiotensin converting enzyme inhibitor, angiotensin receptor II blocker, calcium channel blocker, alpha blocker, thiazide diuretics, hydralazine). Other diabetes drugs refer to a pre-baseline prescription for a sulfonylurea, GLP-1 agonist or baseline insulin. Anticoagulant refers to a pre-baseline prescription for warfarin or a direct oral anticoagulant. Antiplatelet refers to a pre-baseline prescription for clopidogrel, ticagrelor, prasugrel or dipyridamole. Cholesterol consists of both low-density and high-density lipoprotein measurements. Socioeconomic status is defined at the level of the GP practice using the UK national index of multiple deprivation quintile. Ethnicity is categorised as either: white, black, Asian, mixed or other and is further defined in supplement three. Comorbidities refer to a set of individual conditions, each defined separately rather than as a grouped category. These conditions were: heart failure, significant mental illness (schizophrenia, bipolar affective disorder, depression), chronic obstructive pulmonary disease, rheumatoid arthritis, asthma, epilepsy, dementia, inflammatory bowel disease, liver disease.*

2.     	Covariate definitions

Table outlining definitions of variables used in the study. Full repository, including read codes, available online at: <https://tinyurl.com/ac9pxrb>. Comorbidities considered as confounders were selected based on clinical domain knowledge.

**Supplementary table 1:** Covariate definitions

| **Covariate** | **Definition** |
| --- | --- |
| **Demographics** | |
| Age | Age at cohort entry (years) |
| Sex | Recorded sex (male, female) |
| Ethnicity | Recorded ethnicity re-categorised according to definitions in supplementary material 3. |
| Smoking status | Most recent smoking status measurement – categorised as non-smoker, ex-smoker or current smoker. |
| **Prescribing – defined as having a prescribed agent for the following drugs at any time before or on day of cohort entry** | |
| DPP4-i | Alogliptin, linagliptin, sitagliptin, saxagliptin, vildagliptin |
| Baseline metformin prescription | Metformin monotherapy – including modified release formulations |
| Baseline anti-hypertensive agent | Angiotensin converting enzyme inhibitor, angiotensin receptor II blocker, calcium channel blocker, alpha blocker, thiazide diuretics, hydralazine |
| Baseline lipid lowering agent | Statins (simvastatin, rosuvastatin, fluvastatin, atorvastatin, pravastatin) ezetimibe, fibrates |
| Baseline sulfonylurea use | Glibenclamide, gliclazide, glipizide, glimperide, tolbutamide |
| Baseline GLP-1 therapy use | Semaglutide, exenatide, liraglutide, dulaglutide, lixisenatide |
| Baseline insulin use | Defined on the basis of insulin brands. |
| Baseline anticoagulant use | Warfarin and direct oral anticoagulants (apixaban, dabigatran, rivaroxaban, edoxaban) |
| Baseline antiplatelet medication use | Clopidogrel, ticagrelor, prasugrel, dipyridamole |
| **Clinical measurements – defined as measured value within a set window prior to cohort entry** | |
| Glycated haemoglobin A1c (HbA1c) – marker of diabetes severity (mmol/mol) | Most recent measurement within 180 days of cohort entry. |
| Body mass index (BMI) – kg/m^2^ | Most recent measurement within 540 days of cohort entry |
| LDL cholesterol (mmol/L) | Most recent measurement within 540 days of cohort entry |
| HDL cholesterol (mmol/L) | Most recent measurement within 540 days of cohort entry |
| Systolic blood pressure (mmHg) | Most recent measurement within 540 days of cohort entry |
| Estimated glomerular filtration rate (eGFR) -ml/min/1.73m^2^ | Most recent creatinine measurement within 540 days of cohort entry, with eGFR estimated using the National Kidney Foundation recommended CKD EPI 2021 formula – with no adjustment made for ethnicity as this data was missing in a substantial number of people. |
| **Comorbidities – defined as presence of at least one coded diagnosis at any time point before or on date of cohort entry** | |
| Cardiovascular disease | Adapted from HDR UK phenotype library PH894:  <https://phenotypes.healthdatagateway.org/phenotypes/PH894/version/1867/detail/> |
| Type 2 diabetes | Adapted from HDR UK phenotype library PH152 and PH718:  <https://phenotypes.healthdatagateway.org/phenotypes/PH152/version/304/detail/>  <https://phenotypes.healthdatagateway.org/phenotypes/PH718/version/304/detail/> |
| Heart failure | Adapted from HDR UK phenotype library PH182:  <https://phenotypes.healthdatagateway.org/phenotypes/PH182/version/304/detail/> |
| Significant mental illness (schizophrenia, bipolar affective disorder, depression) | Adapted from HDR UK phenotype library PH285, PH149, PH38:  <https://phenotypes.healthdatagateway.org/phenotypes/PH285/version/304/detail/>  <https://phenotypes.healthdatagateway.org/phenotypes/PH149/version/304/detail/>  <https://phenotypes.healthdatagateway.org/phenotypes/PH38/version/304/detail/> |
| Chronic obstructive pulmonary disease | Adapted from HDR UK phenotype library PH43:  <https://phenotypes.healthdatagateway.org/phenotypes/PH43/version/304/detail/> |
| Rheumatoid arthritis | Adapted from HDR UK phenotype library PH80:  <https://phenotypes.healthdatagateway.org/phenotypes/PH80/version/304/detail/> |
| Asthma | Adapted from HDR UK phenotype library PH109:  <https://phenotypes.healthdatagateway.org/phenotypes/PH109/version/304/detail/> |
| Epilepsy | Adapted from HDR UK phenotype library PH467:  <https://phenotypes.healthdatagateway.org/phenotypes/PH467/version/304/detail/> |
| Dementia | Adapted from HDR UK phenotype library PH473:  <https://phenotypes.healthdatagateway.org/phenotypes/PH473/version/304/detail/> |
| Inflammatory bowel disease | At any point before or on date of cohort entry.  Adapted from HDR UK phenotype library PH596:  <https://phenotypes.healthdatagateway.org/phenotypes/PH596/version/304/detail/> |
| Liver disease | At any point before or on date of cohort entry.  Adapted from HDR UK phenotype library PH1085:  <https://phenotypes.healthdatagateway.org/phenotypes/PH1085/version/304/detail/> |

3.     	Ethnicity

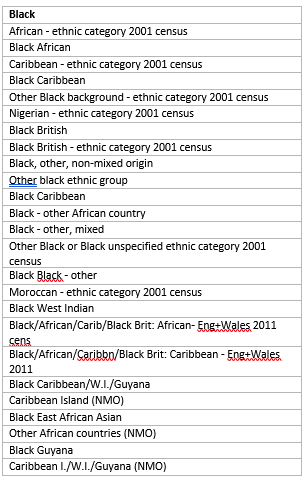
Ethnicity was re-coded to align with the most recent UK census classification of ethnicity (White, Black, Asian, Other, mixed).^1^

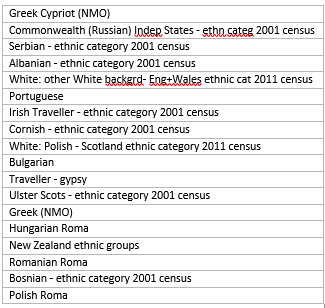

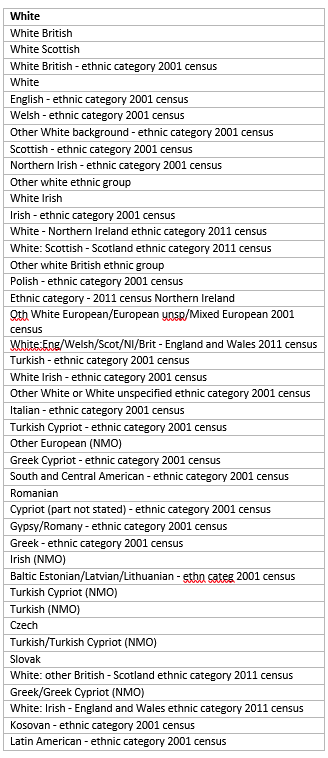

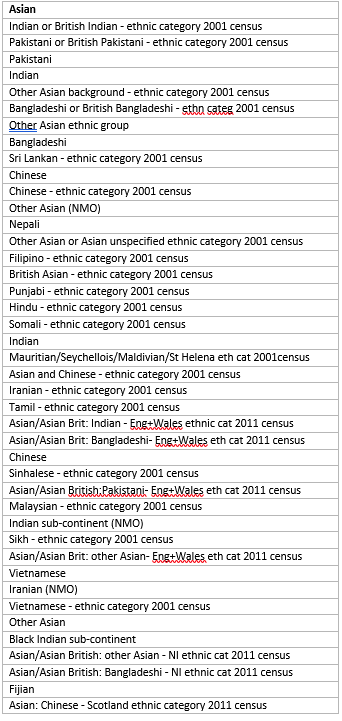

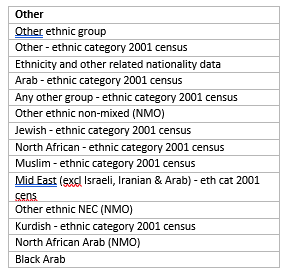

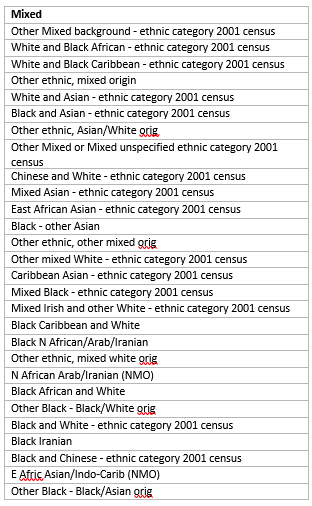

4.     	Dataset construction

This study uses data from the health improving network (THIN), which is a representative UK primary care database that captures non-identifiable electronic health record data for 20 million people.^2,3^ THIN was not linked with any other data source.

An extract from the THIN database was obtained containing data for people who had at least one prescription for a study-relevant drug (SGLT2i: empagliflozin; DPP-4i: alogliptin, linagliptin, sitagliptin, saxagliptin and vildagliptin) between 1st January 2014 and 31st December 2022.

Data analysis was conducted in the University College London data safe haven. This is a secure, virtual analytic platform that conforms to national and NHS Digital information governance policies. All data will be stored and analysed within this environment, with exports of the results of final analysis being authorised by the data manager to ensure no breech of confidential information. Data management was conducted in python 3.9.16 and analysis was conducted in Stata 18 and R version 4.4.1. Code notebooks are available online: <https://github.com/dkdryan/empa_reg_tte>.

The data cleaning pipeline first identified spurious dates for treatment initiation and event dates (i.e. recorded death dates in the future, people who had a last contact date before or on the date of cohort entry). These were removed from the dataset, with numbers retained in the flowchart (figure 1, main text and figure 2, main text) to demonstrate how many people were excluded due to these administrative criteria.

Secondly, the distribution of laboratory and clinical measurements (glycated haemoglobin A1c, body mass index, systolic blood pressure, low-density lipoproteins and high-density lipoproteins) wasassessed. Values beyond the maximum and minimum limits of the distribution within the UK biobank were deemed unreliable and set to missing.^4^ These limits are defined below:

**Supplementary table 2:** Parameters to define extreme values were based on the maximum and minimum values for the distribution of the clinical measurement in the UK biobank

| **Clinical variable (unit)** | **Lower limit** | **Upper limit** | **Reference** |
| --- | --- | --- | --- |
| HbA1c (mmol/mol) | 15 | 55 | <https://biobank.ndph.ox.ac.uk/showcase/field.cgi?id=30750> |
| BMI (kg/m^2^) | 10 | 80 | <https://biobank.ndph.ox.ac.uk/ukb/field.cgi?id=21001> |
| Creatinine (mmol/L) | 10 | 1500 | <https://biobank.ctsu.ox.ac.uk/ukb/field.cgi?id=30700> |
| Aspartate aminotransferase (I/U) | 3 | 950 | <https://biobank.ndph.ox.ac.uk/showcase/field.cgi?id=30650> |
| Alanine aminotransferase (I/U) | 3 | 500 | <https://biobank.ndph.ox.ac.uk/showcase/field.cgi?id=30620> |
| Alkaline phosphatase (I/U) | 8 | 1500 | <https://biobank.ndph.ox.ac.uk/showcase/field.cgi?id=30610> |
| LDL cholesterol (mmol/L) | 0.2 | 10 | <https://biobank.ndph.ox.ac.uk/ukb/field.cgi?id=30780> |
| HDL cholesterol (mmol/L) | 0.2 | 5 | <https://biobank.ndph.ox.ac.uk/showcase/field.cgi?id=30760> |
| Systolic blood pressure (mmHg) | 50 | 270 | <https://biobank.ctsu.ox.ac.uk/crystal/field.cgi?id=4080> |

5.     	Mapping the EMPA-REG RCT eligibility criteria to real-world populations

| **Criteria** | **EMPA-REG Outcome RCT** | **Trial Emulation: RCT Eligible Definition** |
| --- | --- | --- |
| **Inclusion** | People aged 18 years or older with a T2DM diagnosis | People aged 18 years or older with a coded T2DM diagnosis, registered on database for a year or more |
|  | Stable T2DM management - defined as either: (i) no glucose-lowering agents and a HbA1c between 53 and 75mmol/mol 12 weeks prior to the study or (ii) no change in glucose-lowering agents and a HbA1c between 53 - 86mmol/mol 12 weeks prior to the study | Commenced on empagliflozin or DPP-4i for the first time between 1st January 2014 and 31st December 2022. HbA1c 53 – 86mmol/mol within 180 days of cohort entry. If missing – person was not excluded |
|  | High risk of CVD – defined as: MI, multi-vessel coronary artery disease, single-vessel CAD with a positive stress test or UA hospitalisation in previous year, UA and prior evidence of CAD, occlusive peripheral artery disease | High risk of CVD – defined as presence of ACS, ischaemic heart disease, cerebrovascular accident (stroke/TIA), peripheral artery disease prior to, or on date of cohort entry |
|  | Baseline BMI ≤45 kg/m2 | Recorded BMI ≤ 45kg/m2 within 540 days of cohort entry. If missing – person was not excluded |
| **Exclusion** | Uncontrolled hyperglycaemia with fasting glucose >13.3 mmol/mol | Not possible to map |
|  | Liver disease with liver function tests > 3x upper limit of normal | Patients with a liver function test > 3x upper limit of normal within 2 months of cohort entry |
|  | Planned cardiovascular surgery or angioplasty in 3 months | Not possible to map |
|  | People who had an ACS or stroke/TIA event in the past 2 months | People who had a coded diagnosis for ACS, stroke or TIA in the 2 months preceding cohort entry |
|  | eGFR <30 mL/min/1.73m2 | Participants whose last recorded eGFR was < 30 ml/min/1.73m2, with a limit set at 540 days. If missing – person was not excluded |
|  | Prior surgery for chronic malabsorption (e.g. bariatric) in prior 2 years | People who had bariatric surgery within the last 2 years |
|  | Red-blood cells disorders | People with a malaria diagnosis in past 5 years |
|  | Cancer within the preceding 5 years | People who had a cancer diagnosis or treatment in past 5 years |
|  | Anti-obesity drug treatment in prior 3 months | Treatment with orlistat in 3 months prior to cohort entry. A |
|  | Systemic steroids within the preceding 6 weeks | People prescribed a systemic steroid used within 6 weeks of cohort entry |
|  | Change in thyroid hormone dosage in prior 6 weeks | People prescribed a change in dose of levothyroxine within 6 weeks of cohort entry |
|  | Nursing, pregnant, or child-bearing aged women not using acceptable method of birth control or refusing pregnancy testing; Alcohol or drug abuse | People who had a pregnancy within 12 months prior to cohort entry |
|  | Alcohol or drug abuse | People with a coded diagnosis for significant drug or alcohol misuse in the three months prior to cohort entry |
|  | Not excluded in the EMPA-REG RCT | People who ever had a coded diagnosis for ketoacidosis or pancreatitis at any point prior to cohort entry |

| **Supplementary table 3:** Comparison of eligibility criteria for the EMPA-REG RCT and the trial emulation | | |
| --- | --- | --- |
| **Criteria** | **EMPA-REG RCT** | **Trial Emulation: RCT Eligible Definition** |
| **Inclusion** | People aged 18 years or older with a type 2 diabetes mellitus (T2DM) diagnosis | People aged 18 years or older with a coded T2DM diagnosis, registered on database for a year or more |
|  | Stable T2DM management - defined as either: (i) no glucose-lowering agents and a HbA1c between 53 and 75mmol/mol 12 weeks prior to the study or (ii) no change in glucose-lowering agents and a HbA1c between 53 - 86mmol/mol 12 weeks prior to the study | Commenced on empagliflozin or DPP-4i for the first time between 1st January 2014 and 31st December 2022. HbA1c 53 – 86mmol/mol within 180 days of cohort entry. If missing – a person was not excluded |
|  | High risk of cardiovascular disease – defined as: myocardial infarction, multi-vessel coronary artery disease, single-vessel coronary artery disease with a positive stress test or unstable angina hospitalisation in previous year, unstable angina and prior evidence of coronary artery disease, occlusive peripheral artery disease | High risk of cardiovascular disease – defined as presence of acute coronary syndrome, ischaemic heart disease, cerebrovascular accident (stroke/transient ischaemic attack), peripheral artery disease prior to, or on date of cohort entry |
|  | Baseline BMI ≤45 kg/m2 | Recorded BMI ≤ 45kg/m2 within 540 days of cohort entry. If missing – person was not excluded |
| **Exclusion** | Uncontrolled hyperglycaemia with fasting glucose >13.3 mmol/mol | Not possible to map |
|  | Liver disease with liver function tests > 3x upper limit of normal | Patients with a liver function test > 3x upper limit of normal within 2 months of cohort entry |
|  | Planned cardiovascular surgery or angioplasty in 3 months | Not possible to map |
|  | People who had an acute coronary syndrome or stroke/transient ischaemic attack event in the past 2 months | People who had a coded diagnosis for acute coronary syndrome, stroke or transient ischaemic attack in the 2 months preceding cohort entry |
|  | eGFR <30 mL/min/1.73m^2^ | Participants whose last recorded estimated glomerular filtration rate (eGFR) was < 30 ml/min/1.73m^2^, with a limit set at 540 days. If missing – person was not excluded |
|  | Prior surgery for chronic malabsorption (e.g. bariatric) in prior 2 years | People who had bariatric surgery within the last 2 years |
|  | Red-blood cells disorders | People with a malaria diagnosis in past 5 years |
|  | Cancer within the preceding 5 years | People who had a cancer diagnosis or treatment in past 5 years |
|  | Anti-obesity drug treatment in prior 3 months | Treatment with orlistat in 3 months prior to cohort entry. A |
|  | Systemic steroids within the preceding 6 weeks | People prescribed a systemic steroid used within 6 weeks of cohort entry |
|  | Change in thyroid hormone dosage in prior 6 weeks | People prescribed a change in dose of levothyroxine within 6 weeks of cohort entry |
|  | Nursing, pregnant, or child-bearing aged women not using acceptable method of birth control or refusing pregnancy testing. | People who had a pregnancy within 12 months prior to cohort entry |
|  | Alcohol or drug abuse | People with a coded diagnosis for significant drug or alcohol misuse in the three months prior to cohort entry |
|  | Not excluded in the EMPA-REG RCT | People who ever had a coded diagnosis for ketoacidosis or pancreatitis at any point prior to cohort entry |

6.     	Statistical methods

The primary analytical approach was an adjusted Cox proportional hazard model. In addition, we conducted as sensitivity analysis an inverse-probability of treatment weighting (IPTW) of a Cox proportional hazards model. The IPTW was also employed to estimate an average treatment effect and an average treatment effect in the treated, as measured using a HR. The adjusted Cox proportional hazard model was selected for the primary analysis as it estimates a conditional hazards ratio, which mirrors most faithfully the conditional hazard ratio of the RCT. In addition, IPTW can have issues with lower power. We acknowledge that the adjusted HRs estimated in the RCT and in the trial emulation are conditional on a different set of covariates and therefore not strictly comparable, but do not consider this to be a significant barrier to comparison here.

**Estimating the hazard ratio: Adjusted Cox proportional hazards model**

The adjusted Cox proportional hazards model included treatment status (empagliflozin vs DPP-4i) and all covariates defined in the study DAG (supplementary material 1). To assess the functional form of included continuous covariates, Martingale residuals were used, with continuous variables tested for inclusion in untransformed, quadratic, log and spline (3 knots) forms. No interactions were included for the main analysis. The functional form investigations were performed based on the complete case data. The final functional form of the variables was determined by visually inspecting loess plots of Martingale residuals against the continuous variables to determine appropriate functional form The process resulted in inclusion of both linear and squared terms for two of the continuous covariates: LDL cholesterol and eGFR, and in HDL being log transformed. All other continuous covariates were included untransformed.

The model was assessed for the assumption of proportional hazards using scaled Schoenfeld residual plots, plots of the -log cumulative hazard against log time and assessing the hypothesis test for non-zero slope in a generalised linear regression of the scaled Schoenfeld residuals on time. There was no evidence to suggest a major breach of the proportional hazards assumption.

The adjusted Cox model assumes that the censoring time is independent of the event time conditional on the variables included in the model and that the model is correctly specified, including that there are no interactions between treatment and covariates.

**Estimating the hazard ratio: Inverse-probability of treatment weighted (IPTW) Cox proportional hazards model**

The inverse-probability of treatment weighting (IPTW) method is a two-step process which initially estimates the probability of treatment for each individual – that is the conditional probability that an individual was prescribed empagliflozin conditional on their covariates, regardless of their actual treatment.

Each individual is assigned a weight that is the inverse of the estimated probability of their observed treatment given their covariates. More specifically the average treatment effect (ATE) weight W for individual *i* was estimated as being W*_i_* = 1/p(C*_i_*) for those in the empagliflozin group and W*_i_* = 1/1 – p(C*_i_*) for those in the DPP-4i group, where p(C*_i_*) is the estimated probability of being treated with empagliflozin given covariates C*_i._* There was no evidence of extreme weights and good overlap between treatment groups. These inverse probability of treatment weights are then used in a weighted outcome Cox proportional hazards model, with treatment assignment as the only covariate. The weights seek to balance covariates between treatment groups, accounting for baseline imbalance in a non-randomised, observational setting. This results in an estimate of a marginal HR, representing a type of average treatment effect.

We also estimated the average treatment effect in the treated (ATT) using a similar process but with modified weights. For the ATT the weight *W* for individual *i* was estimated as being *W_i_* = 1 for those in the empagliflozin group and *W_i_* = p(C*_i_*)*_/_*1 – p(C*_i_*) for those in the DPP-4i group, where p(C*_i_*) is as defined above.

The conditional treatment probabilities used in the IPTW analyses were derived from a logistic regression model with empagliflozin allocation as the outcome and including all the covariates from the adjusted Cox proportional hazards model. These covariates are related to the exposure and the outcome. No interactions were included. The resulting weights were examined using histograms to investigate whether there were any disproportionately large weights, and the treatment probabilities distribution was compared in the empagliflozin and DPP-4i groups to assess the extent of the overlap.

To assess covariate balance following IPTW weighting, average standardised mean differences between covariates in the weighted treatment groups were estimated and presented on a forest plot to compare covariate balance in weighted populations compared to the raw unweighted population. Standardised mean differences were obtained in five imputed data sets and then combined using Rubin’s rules. The assessment of covariate balance was performed using the *matchthem* and *cobalt* packages in R. Adequate balancing of covariates between treatment groups was defined as a standardised mean difference of less than 10% between groups for a covariate.

Based on the IPTW Cox model, the estimated marginal hazard ratio for all-cause mortality with associated 95% confidence interval was obtained. The standard errors were estimated using the robust sandwich estimator to account for uncertainty introduced by the propensity score estimation. The IPTW analysis assumed independent censoring, proportional hazards assumption and correctly specified propensity score model, but does not make assumptions about treatment-covariate interactions.

**Missing data**

Missing data in confounding variables was described using descriptive statistics to determine patterns of missingness. Variables with missingness had their missing values imputed using multiple imputation by chained equations (MICE), implemented in R with the *mice* package. This method is valid under the assumption that the data are missing at random (MAR). MAR describes a situation where the probability of missingness in the partially observed variable is independent of the value of that variable, conditional on the fully observed variable(s). MICE enables us to use the distribution of the fully observed covariates to estimate multiple possible values for the data points. In addition, it appropriately accounts for the uncertainty in estimation introduced by imputation. It is an iterative process that estimates incomplete variables using separate models, based on complete data and previously imputed data. This is repeated until estimates of imputed values converge. In this procedure, five iterations were used.

The imputation model contained all the variables in the substantive model and the outcome variables, which in a survival context, were the binary outcome (death within follow-up) and the Nelson-Aalen estimator of cumulative hazard at an individual’s observed event or censoring time.^1^ Categorical variables (ethnicity, smoking status) were imputed using a multinomial logistic regression model and continuous variables (BMI, LDL cholesterol, HDL cholesterol, HbA1c) were imputed using a predictive mean matching method.^2^ Predictive mean matching is a method of imputing missing data by matching each missing value with an observed value from a donor with a similar predicted outcome. An advantage of this method is that it prevents the imputed values falling outside plausible range of values.^2^

No interaction terms were present in the substantive model and therefore, the imputation model did not include interaction terms.^2^As derived terms (squared terms for LDL cholesterol and eGFR) were present in the final substantive model, these were passively imputed. This means that, for given covariates *X* and *X^2^*, missing values of *X* are imputed first, and then the *X^2^* values are calculated by squaring the imputed value of *X.* The decision to use passive imputation was based on evidence from simulation studies which suggest that predictive mean model imputation followed by passive imputation of derived variables is the least biased for logistic regression outcome models, compared to other methods such as ‘just another variable’ approaches.

Five imputed datasets were separately generated using this approach. The propensity score model was fitted separately in each imputed data set. The final outcome model was applied separately to each of the five imputed datasets and pooled estimates for the HR and SE were derived using Rubin’s rules. For the IPTW model, the treatment effect estimates from each imputed dataset were combined to obtain the pooled estimate. This method, known as MIte (multiple imputation treatment effect), has been shown to provide unbiased estimates in studies involving MI for IPTW analysis.^3^

**G-computation methods to estimate marginal risk of mortality**

The marginal risk difference in all-cause mortality at three years was estimated using a g-computation approach, which allowed for the comparison of the average risks between treatment groups while controlling for confounding variables. This procedure was as follows:

1. The adjusted Cox model were used to estimate the survival probability for each individual at three years under both treatment assignments (empagliflozin or DPP-4i) and given their values of the covariates included in the model.
2. This was converted to an individual’s three-year risk of all-cause mortality by subtracting survival probability from 1.
3. For each individual, the difference between the risk of all-cause mortality under both treatment assignments at three years was estimated, giving and individualized risk difference.
4. The average difference in risk under empagliflozin vs DPP-4i was calculated across the entire cohort by averaging the individualized differences. This represented the marginal risk difference in all-cause mortality at three years.

Confidence intervals for the marginal risk difference were generated using a bootstrap procedure. Since the data required methods for handling missing data (via multiple imputation by chained equations, MICE) alongside a bootstrap procedure to generate confidence intervals for the g-computation method, a combined bootstrap-multiple imputation (Boot MI) method was employed. This was based on recommendations from Bartlett and Von Hippel and was implemented using the *bootImpute* package in R.^4,5^

As per guidance from Bartlett and Von Hippel, 20 bootstrap datasets were generated, each one was imputed twice.^4,5^ Within each fully imputed bootstrap sample, the marginal risk difference in all-cause mortality at three years was estimated. Von Hippel’s Boot-MI method was then used to pool estimates and determine standard errors from the multiply imputed bootstrapped samples.^4,5^ The effect estimate was determined by first finding the average of the estimates obtained from each of the imputed datasets and then taking the overall average across all bootstrap samples. The standard error was derived using formulas that considered both bootstrap and imputation variability in a computationally efficient manner.

**Agreement metrics**

The standardised difference between the hazard ratio estimated from the RCT and that from the emulated study can be calculated using the following formula:

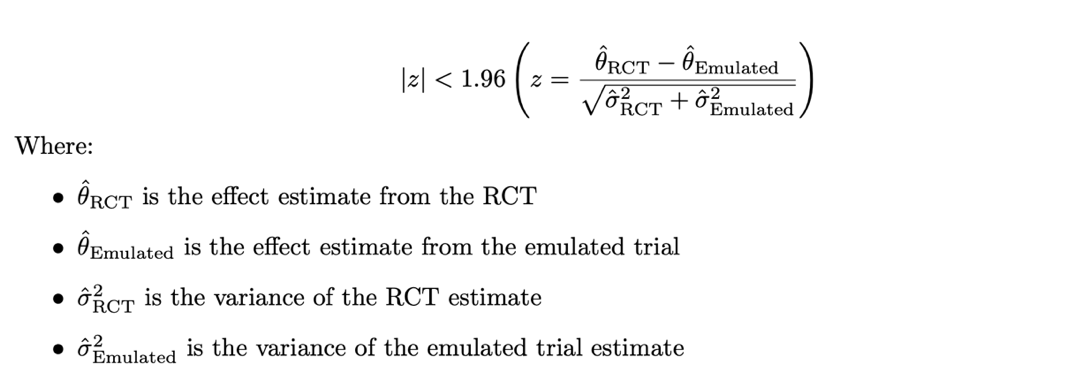

The estimated z-value is then compared to standard normal distribution to derive a p-value. A hypothesis test can then be conducted with the null hypothesis being that no difference exists between the estimate from the RCT and the emulated trial.

**Assumptions for causal inference**

The following assumptions are necessary for a causal interpretation of our results:

1.         	Correctly specified models – this applies to the propensity score, multiple imputation and outcome models.

2.         	Positivity – there is a non-zero probability of receiving either treatment for every combination of exposure and confounder values present in the population.

3.         	Conditional exchangeability – no unmeasured confounding.

4.         	No interference – an individual’s potential outcome is not influenced by treatment received by another individual.

5.         	Consistency – the outcome observed for people under a specific treatment strategy (empagliflozin or DPP-4i) is equal to the potential outcome that person would experience if they had been assigned the same treatment strategy.

**Quantitative bias assessment: E-value estimation**

The E-value was estimated using the following formula:

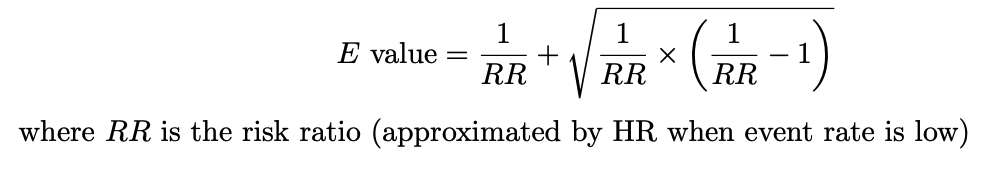

**Interaction between RCT eligibility and treatment effect**

The adjusted Cox proportional hazards model was amended to test for an interaction effect between RCT eligibility and empagliflozin. This enables an assessment of whether there is a significant difference in the treatment effect of empagliflozin on all-cause mortality between people who were RCT eligible versus those who were not RCT eligible. RCT eligibility was defined as any person who would have met the eligibility criteria for the RCT, based on the definitions in supplementary table 3. The indicator of RCT eligibility and an interaction term between RCT eligibility and empagliflozin were added to the model. No other changes were made to the functional form of the model. The hazard ratio, 95% confidence interval and p-value for the interaction term was obtained from the adjusted Cox proportional hazard model. A hypothesis test for an interaction effect was conducted with the null hypothesis being that there was no interaction between treatment assignment and RCT eligibility.

7.	Missing data

**Supplementary table 4:** Proportion of missing values in the trial emulation

|  | **Trial emulation population (N = 62503)** |
| --- | --- |
| Age (years) | - |
| Male (N, %) | - |
| Ethnicity (N, %) | 35739 (52.2%) |
| Smoking status (N, %) | 1281 (2.0%) |
| BMI (kg/m^2^) | 8545 (13.7%) |
| HbA1c (mmol/mol) | 7847 (12.6%) |
| Systolic blood pressure (mmHg) | 3596 (5.8%) |
| Baseline eGFR (ml/min/1.73m^2^) | 2451 (3.9%) |
| Baseline LDL cholesterol (mg/dL) | 14227 (22.8%) |
| Baseline HDL cholesterol (mg/dL) | 7712 (12.3%) |

***Supplementary table 4:*** *Table describing the proportion of missing values for each covariate in the trial and extended trial emulations. BMI: body mass index, HbA1c: glycated haemoglobin, eGFR: estimated glomerular filtration rate, LDL: low-density lipoprotein, HDL: high-density lipoprotein.*

8.     	Demographics according to treatment randomisation/allocation

| **Supplementary table 5:** Real-world and RCT populations according to treatment groups | | | | |
| --- | --- | --- | --- | --- |
|  | **EMPA-REG RCT population (N = 7020)** | | **Trial emulation (N = 62503)** | |
| **Cohort (N, % of population)** | Empagliflozin  (N = 4687; 66.7%) | Placebo  (N = 2333; 33.3%) | Empagliflozin  (N = 13239; 21.2%) | DPP-4i  (N = 49264; 78.8%) |
| Age (years) | 63.1 (8.6) | 63.2 (8.8) | 58.7 (11.6) | 65.4 (13.2) |
| Female (N, %) | 1, 351 (28.9%) | 653 (28.0%) | 5321 (43.1%) | 21214 (40.2%) |
| Ethnicity (N, %)              Missing              White              Asian              Black              Other              Mixed | 41 (0.9)  3, 403 (72.6)  1, 006 (21.5)  237 (5.1)  -  - | 24 (1.0)  1, 678 (71.9)  511 (21.9)  120 (5.1)  -  - | 7390 (55.8%)  5158 (39.0%)  438 (3.3%)  119 (0.9%)  79 (0.6%)  55 (0.4%) | 28, 349 (57.5%)  17, 491(35.5%)  2, 252 (4.6%)  700 (1.4%)  277 (0.6%)  195 (0.4%) |
| Smoking status (N, %)              Current              Ex-smoker              Non-smoker              Missing | -  -  -  - | -  -  -  - | 2228 (16.8%)  5718 (43.2%)  5131 (38.8%)  162 (1.2%) | 6821 (13.8%)  22691 (46.1%)  18633 (37.8%)  1119 (2.3%) |
| Comorbidities (N, %)              Cardiovascular disease              Heart failure              Recent cancer diagnosis/treatment              Chronic obstructive pulmonary disease              Dementia              Epilepsy              IBD              Liver disease              Significant mental illness              Rheumatoid arthritis | -  -  -  -  -  -  -  -  -  - | -  -  -  -  -  -  -  -  -  - | 3424 (25.7%)  672 (5.1%)  389 (2.9%)  831 (6.3%)  50 (0.4%)  252 (1.9%)  212 (1.6%)  352 (2.7%)  4548 (34.4%)  176 (1.3%) | 15407 (31.3%)  3117 (6.3%)  2412 (4.9%)  3673 (7.5%)  1172 (2.4%)  900 (1.8%) 944 (1.9%)  1119 (2.3%)  14288 (29.0%)  846 (1.7%)    *Table continued…* |
| **Supplementary table 5 continued:** Real-world and RCT populations according to treatment groups | | | | |
|  | **EMPA-REG RCT population (N = 7, 020)** | | **Trial emulation (N = 62, 503)** | |
| **Cohort (N, % of population)** | Empagliflozin  (N = 4687; 66.7%) | Placebo  (N = 2333; 33.3%) | Empagliflozin  (N = 13239; 21.2%) | DPP-4i  (N = 49264; 78.8%) |
| BMI (kg/m^2^) | 30.6 (5.3) | 30.7 (5.2) | 34.8 (7.3) | 31.9 (6.7) |
| HbA1c (mmol/mol) | 64 (9.4) | 64 (9.3) | 76.6 (18.3) | 73.7 (17.6) |
| Systolic blood pressure (mmHg) | 135.3 (16.9) | 135.8 (17.2) | 134.5 (15.1) | 133.2 (15.0) |
| Baseline eGFR (ml/min/1.73m^2^) | 74.2 (21.6) | 73.8 (21.1) | 93.1 (17.0) | 80.2 (24.6) |
| Baseline LDL cholesterol (mmol/L) | 2.62 (0.9) | 2.59 (0.9) | 2.7 (1.2) | 2.5 (1.1) |
| Baseline HDL cholesterol (mmol/L) | 1.7 (0.3) | 1.27 (0.3) | 1.1 (0.3) | 1.2 (0.3) |
| Co-prescribed drugs (N, %)  Metformin  Sulfonylurea  GLP1RA  Insulin  Statin/lipid lowering agent  Anti-hypertensive agent  Anticoagulant  Antiplatelet | 3459 (73.8%)  2014 (43.0%)  70 (3.0%)  2252 (48.0%)  3820 (81.5%)  4446 (94.9%)  -  - | 1734 (74.3%)  992 (42.5%)  126 (2.7%)  1135 (48.6%)  1864 (79.9%)  2221 (95.2%)  -  - | 6940 (52.4%)  2965 (22.4%)  965 (7.3%)  1711 (12.9%)  7063 (53.4%)  6266 (47.3%)  564 (4.3%)  876 (6.6%) | 34457 (70.0%)  18001 (36.5%)  1003 (2.0%)  3742 (7.6%)  34800 (70.6%)  31034 (63.0%)  3856 (7.8%)  5307 (10.8%) |

***Supplementary table 5:*** *Table comparing the trial emulation population with the original RCT population, according to treatment assignment (empagliflozin vs. DPP-4i). Data presented as mean (SD) or count (column percentage) for consistency with trial reporting. *Standard deviation not provided for HbA1c mmol/mol unit. Drug and comorbidity definitions available in supplementary material 1 – 2. BMI: body mass index, HbA1c: glycated haemoglobin A1c, eGFR: estimated glomerular filtration rate, LDL: low density lipoprotein, HDL: high density lipoprotein, GLP1RA: GLP-1 receptor agonist. IBD: inflammatory bowel disease, COPD: chronic obstructive pulmonary disease.*

| **Supplementary table 6:** Demographics of the RCT eligible and RCT ineligible real-world populations | | |
| --- | --- | --- |
|  | **RCT eligible population**  **(N = 12970)** | **RCT ineligible population**  **(N = 49, 533)** |
| Allocated to Empagliflozin (N, %) | 2228 (17.2%) | 10742 (21.7%) |
| Age (years) | 70.8 (10.6) | 62.2 (13.2) |
| Female (N, %) | 4561 (35.2%) | 21974 (44.4%) |
| Ethnicity (N, %)                Missing                White                Asian                Black                Other                Mixed | 7752 (60.0%)  4673 (36.0%)  400 (3.1%)  76 (0.6%)  48 (0.4%)  21 (0.2%) | 27987 (56.5%)  17976 (36.3%)  2290 (4.6%)  743 (1.5%)  308 (0.6%)  229 (0.5%) |
| Smoking status (N, %)                Current                Ex-smoker                Non-smoker                Missing | 1749 (13.5%)  7146 (55.1%)  3835 (30.0%)  240 (1.9%) | 7300 (14.7%)  21263 (42.9%)  19929 (40.2%)  1041 (2.1%) |
| BMI (kg/m^2^) | 31.0 (5.3) | 32.9 (7.2) |
| HbA1c (mmol/mol) | 68.2 (8.6) | 75.8 (19.1) |
| Systolic blood pressure (mmHg) | 133.2 (15.4) | 133.6 (15.0) |
| Baseline eGFR (ml/min/1.73m^2^) | 74.4 (22.4) | 85.23 (23.6) |
| Baseline LDL cholesterol (mg/dL) | 2.3 (1.0) | 2.6 (1.1) |
| Baseline HDL cholesterol (mg/dL) | 1.13 (0.3) | 1.16 (0.3) |
| Co-prescribed medication (N, %)                Metformin                Sulfonylurea                Insulin                GLP1RA                Statin/lipid lowering agent                Anti-hypertensive agent | 9006 (69.4%)  5004 (38.6%)  1396 (10.8%)  389 (3.0%)  10849 (83.6%)  10170 (78.4%) | 32391 (65.4%)  15962 (32.2%)  4057 (8.2%)  1579 (2.5%)  31014 (62.6%)  27130 (54.8%) |
| Comorbidities (N, %)              Cardiovascular disease              Heart failure              Recent cancer diagnosis/treatment              Chronic obstructive pulmonary disease              Dementia              Epilepsy              IBD              Liver disease              Significant mental illness              Rheumatoid arthritis | 12970 (100.0%) 1658 (12.8%) 741 (5.7%)  1477 (11.4%)  452 (3.5%) 318 (2.5%) 275 (2.1%) 313 (2.4%) 3932 (30.3%) 247 (1.9%) | 5861 (11.8%) 2131 (4.3%) 2060 (4.2%)  3027 (6.1%) 770 (1.6%) 834 (1.7%)  881 (1.8%) 1158 (2.3%) 14904 (30.1%) 775 (1.6%) |

***Supplementary table 6:*** *Table comparing the RCT eligible and RCT ineligible real-world populations. Data presented as mean (SD) or count (column percentage) for consistency with trial reporting. Drug and comorbidity definitions available in supplementary material 1 – 2. BMI: body mass index, HbA1c: glycated haemoglobin A1c, eGFR: estimated glomerular filtration rate, LDL: low density lipoprotein, HDL: high density lipoprotein, GLP1RA: GLP-1 receptor agonist. IBD: inflammatory bowel disease, COPD: chronic obstructive pulmonary disease.*

9.	Full results table

**Supplementary table 7:** Full results and agreement metrics

|  | **Results** | | | **Agreement metrics between RCT and real-world analysis** | | |
| --- | --- | --- | --- | --- | --- | --- |
|  | **HR for all-cause mortality (95% CI)** | **3-year risk difference in mortality (95% CI)** | **Number needed to treat (95% CI)** | **Estimate agreement** | **Statistical agreement** | **Standardised difference** |
| **Clinical trial (N = 7001)** | | | | | | |
| EMPA-REG RCT | 0.69 (0.57, 0.82) | Not reported | 38* | - | - | - |
| **Trial emulation (N = 62503)** | | | | | | |
| Unadjusted | 0.42 (0.38, 0.46) | **-** | **-** | ✗ | ✓ | ✗ |
| Adjusted Cox model | 0.76 (0.69, 0.83) | -0.02 (-0.03, -0.01) | 47 (30, 66) | ✓ | ✓ | ✓ |
| **Stratified trial emulation (N = 62503)** | | | | | | |
| RCT eligible population  (N = 12970) | 0.69 (0.56, 0.87) | -0.02 (-0.04, -0.01) | 44 (15, 73) | ✓ | ✓ | ✓ |
| RCT ineligible population  (N = 49533) | 0.78 (0.70, 0.87) | -0.02 (-0.03, -0.01) | 58 (30, 86) | ✓ | ✓ | ✓ |

***Supplementary table 7****: Table describing the estimates (adjusted hazard ratio for all-cause mortality, 3-year risk difference and number needed to treat) and 95% confidence intervals, as well as results of agreement metrics for comparison with the published RCT results. G-computation was not conducted for the complete case analysis. No risk difference was estimated in the RCT. The NNT was provided by a post-hoc analysis of the RCT, but the 95% CI was not provided. The formal benchmark study is the comparison between RCT eligible population and the EMPA-REG RCT published results.*

10.	E-value: quantitative bias assessment results

We assessed bias using E-values, which quantify the minimum strength of association a binary unmeasured confounder would need to have with both the treatment and the outcome to fully explain away the observed association,[28] and can give insight into whether the findings are robust to potential unmeasured confounding.

The E-value for treatment assignment was risk ratio 1.98 (95% CI 1.70 – 2.27, supplementary material 10). This suggests that an unmeasured confounder would need to be strongly associated with both treatment and outcome (risk ratio ≥ 1.98) to fully explain the observed association. It is unlikely that a potential confounder with such a strong association with both treatment and the outcome would have been overlooked in the study. This supports the robustness of the treatment–outcome association against unmeasured confounding

**Supplementary figure 2:** Forest plot showing the E-value (a risk ratio) and the hazard ratios (approximately risk ratios) for confounders within the study.

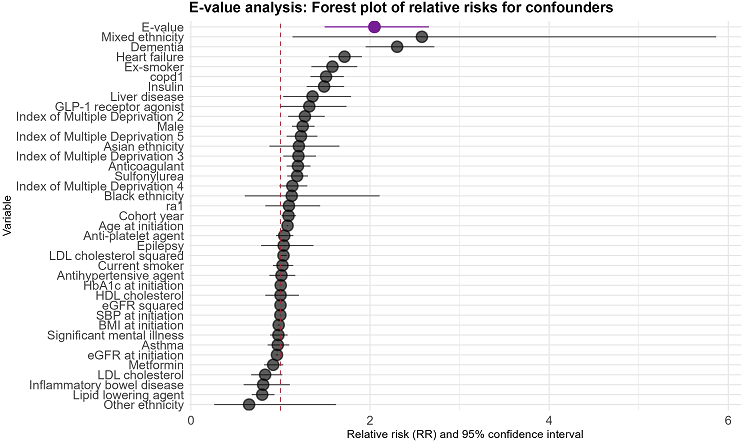

Risk ratio and 95% confidence interval

***Supplementary figure 2:*** *E-value analysis showing the magnitude and 95% confidence interval of a potential unmeasured binary confounder (denoted in purple) that would need to be present to explain away the treatment effect estimated in the adjusted Cox model. Also shown are HRs for confounders from the* *adjusted Cox proportional hazard model, which approximate the risk ratio. COPD: chronic obstructive pulmonary disease. GLP1RA: GLP-1 receptor agonists. RA: rheumatoid arthritis. LDL: low-density lipoprotein cholesterol. HDL: high-density lipoprotein cholesterol.*

11.	Covariate balance for the IPTW analysis

**Supplementary figure 3a:** Covariate balance plot for the average treatment effect

 
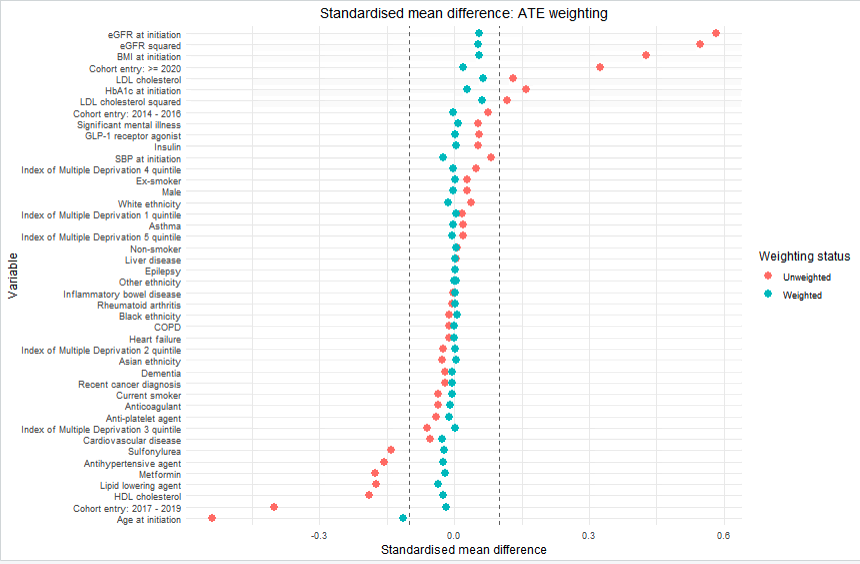

**Supplementary figure 3b:** Covariate balance plots for the average treatment effect in the treated

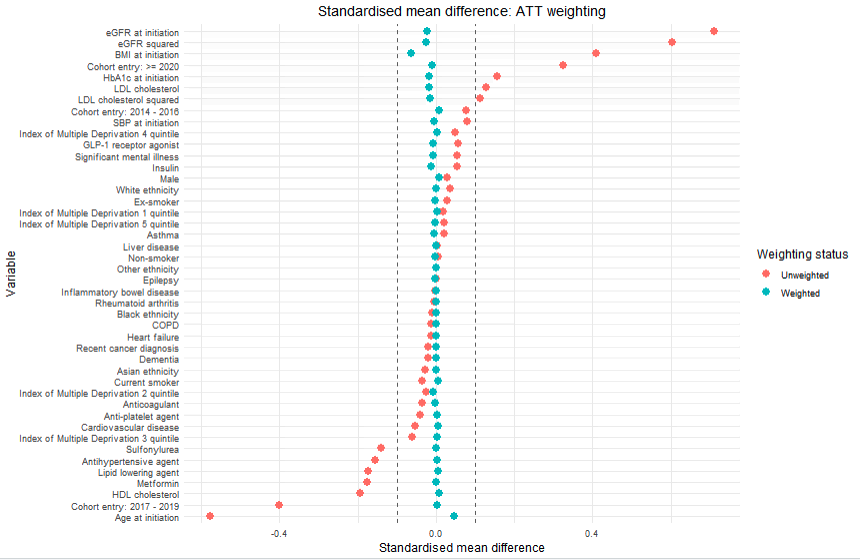

***Supplementary figure 3:*** *Covariate balance plots showing the standardised mean difference between raw and weighted populations after application of the IPTW for the trial emulation using both ATE and ATT weights. Presented standardised mean differences reflect the average across all five multiply imputed datasets. Vertical black lines denote the threshold of acceptable covariate balance ± 0.1. ATE: average treatment effect. ATT: average treatment in the treated population.*

 12.	Average treatment effect and average treatment in the treated estimates

**Supplementary table 8:** IPTW results according to ATE and ATT weights

| **Estimand** | **All-cause mortality adjusted hazard ratio** | **95% confidence interval** |
| --- | --- | --- |
| **Average treatment effect**  **(ATE)** | 0.67 | 0.54 – 0.85 |
| **Average treatment in the treated (ATT)** | 0.74 | 0.66 – 0.82 |
